# Tobacco Use is Related to Parietal-Hippocampal Connectivity in People at Clinical High Risk for Psychosis

**DOI:** 10.64898/2026.05.26.26354136

**Authors:** Yunong Bai, Maxwell J. Roeske, Adam Beermann, Jean Addington, Carrie E. Bearden, Kristin Cadenhead, Tyrone D. Cannon, Ricardo E Carrión, Barbara Cornblatt, Matcheri Keshavan, Daniel H. Mathalon, Diana O. Perkins, Larry Seidman, William S. Stone, Ming T. Tsuang, Elaine F. Walker, Scott Woods, Roscoe O. Brady, Heather Burrell Ward

**Affiliations:** Department of Psychiatry and Behavioral Sciences, Vanderbilt University Medical Center; Department of Psychiatry, Beth Israel Deaconess Medical Center and Harvard Medical School, Boston, MA; Department of Psychiatry, Hotchkiss Brain Institute, University of Calgary, Alberta, Canada; Semel Institute for Neuroscience and Human Behavior, Departments of Psychiatry and Behavioral Sciences and Psychology, University of California, Los Angeles, Los Angeles, CA, USA; Department of Psychiatry, University of California, San Diego, La Jolla, CA, USA; Department of Psychology and Psychiatry, Yale University, New Haven, CT, USA; Department of Psychiatry, Zucker Hillside Hospital and Donald and Barbara Zucker School of Medicine at Hofstra/Northwell, Glen Oaks, NY, USA; Department of Psychiatry and Behavioral Sciences, University of California, San Francisco, San Francisco, CA, USA; Veterans Affairs San Francisco Health Care System, San Francisco, CA, USA; Department of Psychiatry, University of North Carolina at Chapel Hill, Chapel Hill, NC, USA; Department of Psychology, Emory University, Atlanta, GA, USA; Department of Psychiatry, Yale University, New Haven, CT, USA

**Author notes:** <u>Corresponding Authors:</u> Heather Burrell Ward, MD, Department of Psychiatry and Behavioral Sciences Vanderbilt University Medical Center, 1601 23^rd^ Ave S Nashville, TN 37212.

## Abstract

**Background:** Tobacco use is prevalent in clinical high risk for psychosis (CHR-P) population and has widespread negative health consequences, but understanding of its neural substrates is limited. Abnormal default mode network (DMN) may underlie tobacco dependence in CHR-P. We investigated how tobacco use relates to DMN connectivity and how CHR-P status impacts this relationship.

**Methods:** We used baseline substance use and resting-state functional magnetic resonance imaging data from the North American Prodrome Longitudinal Study (NAPLS2; CHR-P: n=211, mean age 19.2, 37.9% female; healthy control: n=132, mean age 19.9, 47.7% female). Voxel-wise connectivity was calculated from the left lateral parietal (LLP) node of the DMN to the rest of the brain. We regressed LLP-brainwide connectivity against tobacco use frequency in the past month to generate a spatial map of how connectivity relates to current tobacco use.

**Results:** Brainwide connectivity analysis identified two clusters in R hippocampus (peak voxel at MNI [+30,-12,-27]) and in L parahippocampus (peak voxel at MNI [-27,-27,-27]), where higher LLP-cluster connectivity was associated with more frequent tobacco use. LLP - R hippocampus connectivity was higher in current tobacco users compared to non-tobacco users (t=-3.5466, df=101.88, p=0.0006), and higher in CHR-P than controls (t=-2.8651, df=279.47, p=0.0049). Among current tobacco users, there was a significant tobacco-by-diagnosis interaction on LLP - R hippocampus connectivity (estimate=0.306, SE=0.149, t=2.051, p=0.045) such that heavier tobacco use predicted hyperconnectivity only in CHR.

**Conclusions:** More frequent tobacco use was associated with higher DMN-hippocampal connectivity in both CHR-P and controls. CHR-P diagnosis enhanced this relationship.

## Introduction

Individuals with schizophrenia (SZ) have a 20-year life expectancy gap compared to the general population, driven by a combination of preventable physical health issues, fragmented healthcare access, and suicide.^1^ Tobacco use is the leading preventable cause of early mortality in SZ^2^. Individuals at clinical high risk for psychosis (CHR-P) experience attenuated positive symptoms of psychosis, brief psychosis episodes, or have genetic risk for psychosis and demonstrates functional decline^3^. Compared to the general population, tobacco use is more prevalent both in SZ^4,5^ and in clinical high risk for psychosis (CHR-P) population^6^. However, current pharmacologic and neuromodulation treatments for smoking cessation are less effective in psychotic disorders compared to that in the general population^7^. There remains a significant gap in our understanding of what neurobiological mechanisms underlies tobacco dependence in both CHR-P and SZ, and what treatment are more effective.

Numerous studies have used functional neuroimaging to study the neural substrates of tobacco dependence in SZ. A novel link was discovered by an agnostic, data-driven analysis regarding tobacco dependence and altered organization of the default mode network (DMN), a resting-state brain network involved in self-referential thoughts^8^ and linked to cognitive performance^9^. This was a SZ-specific relationship and not observed in smokers without SZ^10^. Other studies also reported that stronger tobacco craving was linked to higher DMN connectivity in SZ^11,12^, and that nicotine administration normalized baseline DMN hyperconnectivity^10^.

Brain connectivity undergoes rapid changes during the early stages of development, including in individuals experiencing psychosis and the CHR-P population^13^. Abnormal DMN connectivity has been observed in CHR-P. CHR-P had higher connectivity between DMN and occipital cortex than the control population^14^. Negative symptom severity in CHR-P correlated with higher resting-state connectivity between DMN and other cortical networks, such as salience network and executive control network^13^. CHR-P also exhibited insufficient DMN suppression while performing working memory tasks^15^. However, a recent study reported that smoking was not associated with DMN connectivity differences in CHR-P^16^, but prior literature has yet to investigate dose-dependent relationships between smoking and network connectivity. Altogether, these studies suggest that the DMN is a potential avenue for studying psychosis-specific neural substrate of tobacco dependence.

Therefore, we set out to investigate how tobacco use relates to resting state functional connectivity from LLP, and how CHR-P diagnosis impacts this relationship. We analyzed baseline data from the North American Prodrome Longitudinal Study (NAPLS2), a dataset of CHR-P and healthy control participants with resting state functional neuroimaging and substance use assessment. Given the involvement of the DMN in both the CHR-P period and tobacco use, we hypothesized that LLP would be hyperconnected with the rest of the DMN with increased tobacco use, and this effect would be enhanced in CHR-P individuals.

## Methods

### Participants

We used data from the North American Prodrome Longitudinal Study (NAPLS2), a large, multisite study of young people identified as being at high risk for developing psychosis. Data from 211 individuals at CHR-P and 132 healthy controls who were recruited to the NAPLS2 were included in the study^17^. Individuals at CHR-P were included if they met the Criteria of Prodromal Syndromes (COPS), based on the Structured Interview for Psychosis-Risk Syndromes (SIPS)^18^. Prior to participation, all participants provided written informed consent in accordance with the institutional review boards of Beth Israel Deaconess Medical Center, Boston, Massachusetts; Emory University, Atlanta, Georgia; University of Calgary, Alberta, Canada; University of California, Los Angeles; University of California, San Diego; The University of North Carolina at Chapel Hill; Yale University, New Haven, Connecticut; and Zucker Hillside Hospital, Queens, New York. See Supplement for details.

### Substance Use Measurement

Current substance use was measured using the Alcohol Use Scale/Drug Use Scale (AUS/DUS), which assesses substance use severity (abstinent, use without impairment, abuse, dependence, dependence with institutionalization) and frequency over the past 30 days for the following substances: tobacco, alcohol, marijuana, cocaine, opiates, phencyclidine (PCP), amphetamines, methylenedioxymethamphetamine (MDMA), gamma-hydroxybutarate, huffing, hallucinogens, and other substances. Tobacco use frequency was assessed over the past 30 days on an ordinal scale, rated in cigarettes per day (0 = no use, 1 = occasionally, 2 = <10 per day, 3 = 11-25 per day, 4 = >25 per day). Current tobacco users were grouped into light-use (tobacco use frequency <10 per day) and heavy-use (tobacco use frequency >10 per day).

### MRI Acquisition

NAPLS2 imaging procedures and assessments were designed in a highly coordinated manner. All procedures were harmonized from conception and design of the study prior to data collection^17^. Imaging was conducted on either Siemens 3.0-T MRI systems (Munich, Germany) or GE 3.0-T MRI systems. Briefly, 1-mm^3^ T1-weighted anatomical scans were acquired, and resting-state functional runs of approximately 5 minutes were acquired from all participants (154 time points, 2-second repetition time, 3-mm^3^ voxels). See Supplement for details.

### MRI Data Processing

All analyses were preprocessed using the Data Processing and Analysis for Brain Imaging toolbox (http://rfmri.org/dpabi)^19^. As a quality control metric, scans that exceeded motion thresholds (>3mm translation or >3 degrees rotation) were discarded. Individual time points with framewise displacement >0.2mm were discarded, and scans with >50% of volumes removed for framewise displacement were discarded. All data were preprocessed to remove motion (24-parameter), CSF signals, white matter signals, and an overall linear trend. A bandpass filter was applied (0.01-0.08 Hz). Data were normalized using the DARTEL toolbox (http://www.neurometrika.org/node/34) into Montreal Neurological Institute (MNI) space and smoothed with an 8-mm full-width half-maximum kernel. Voxels within a pre-defined (MNI) gray matter mask were used for further analysis. Data were resampled into 4mm isotropic resolution prior to further analyses. See Supplement for details.

### Unrestricted, Brain-wide Functional Connectivity Analysis

Brainwide analysis was performed on the entire sample. We calculated voxel-wise connectivity by extracting the time course of the BOLD signal from a 6mm sphere placed at left lateral parietal LLP coordinates (MNI [-46, −66, +30])^20^. Using SPM12 (Statistical Parametric Mapping, http://www.fil.ion.ucl.ac.uk/spm), we regressed the z-transformed Pearson’s correlation coefficient connectivity maps against AUS/DUS tobacco use frequency, while controlling for age, sex assigned at birth, and study site as covariates, to generate spatial maps of how connectivity from the LLP region to the entire brain varied with tobacco use frequency at an uncorrected voxelwise threshold of p<0.001 and cluster-forming threshold of p<0.05.

We used “cluster” to refer to each group of voxels with significant correlation in the generated spatial map, and “peak voxel” to refer to the voxel at the location of maximal connectivity-tobacco use association in the cluster. We calculated region-to-seed connectivity by measuring BOLD correlation between the LLP region and 6mm spheres (seeds) placed at the peak voxel in the cluster using Matlab (R2022b Version 9.13.0 (MathWorks, Inc.)). Using the same procedure, we also calculate region-to-seed connectivity for voxels contralateral to the peak voxel.

### Statistical Analyses

Pearson’s correlation coefficients were used to determine the relationships between functional connectivity and tobacco use frequency. T-tests were used to compare continuous outcomes based on dichotomous variables. Gaussian linear models were used to model connectivity based on tobacco use frequency and based on diagnosis x tobacco use interaction, controlling for age, sex, study site, and cannabis use frequency. Robust standard error was used to obtain statistical inferences of linear models given dataset heteroscedasticity. Estimated marginal means were used to compare predicted averages from linear models. All analyses were conducted in RStudio (Version 4.5.2 (2025-10-32)) using alpha<.05.

## Results

Sample demographics and tobacco use overview have been reported elsewhere.^21^

### Higher LLP – Bilateral Hippocampus Connectivity Correlated with Heavier Tobacco Use

To identify brain regions which connectivity to LLP are correlated with tobacco use frequency, we performed brainwide analysis on the entire sample. We identified two clusters (Figures 1A, 2A): 1) R hippocampus (peak voxel at MNI [+30, −12, −27]) and 2) L parahippocampus (peak voxel at MNI [−27, −27, −27]), where higher LLP-cluster connectivity was associated with more frequent current tobacco use (Figures 1B, 2C).

**Figure 1.**
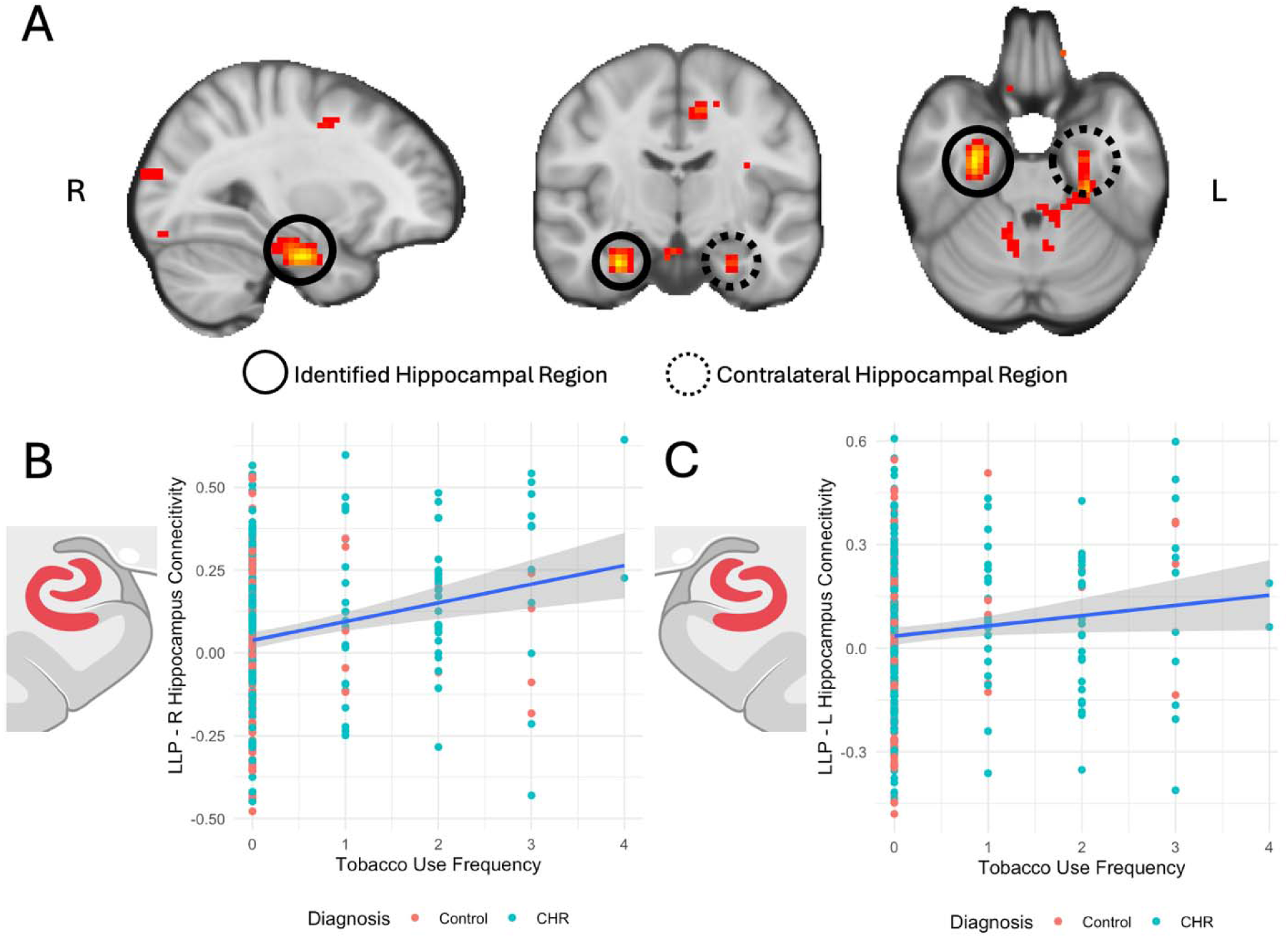
(A) Brain-wide functional connectivity analysis identified regions in the hippocampus whose connectivity to LLP increased with heavier tobacco use. The cluster in R hippocampus (solid circle) reached cluster-level significance and the contralateral cluster (dashed circle) did not. Images were taken at MNI [+30, −12, −27]. (B) Visual representation of the relationship between LLP – R hippocampus connectivity and tobacco use frequency. (C) On the contralateral side, LLP – L hippocampus connectivity was also significantly associated with tobacco use frequency (estimate=0.035, SE=0.016, t=2.175, p=0.031).

**Figure 2.**
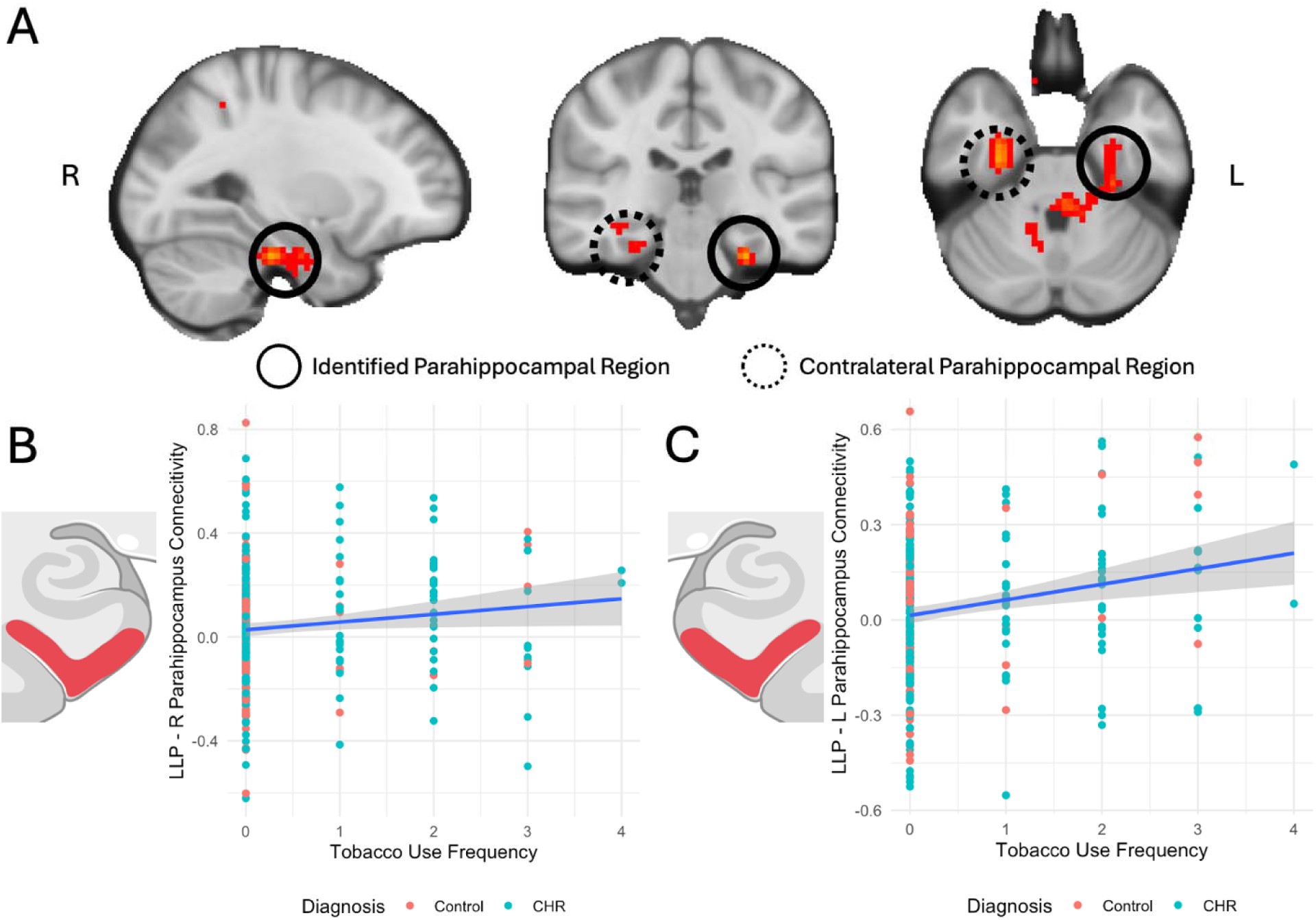
(A) Brain-wide functional connectivity analysis identified regions in the parahippocampus whose connectivity to LLP increased with heavier tobacco use. The cluster in L parahippocampus (solid circle) reached cluster-level significance and the contralateral cluster (dashed circle) did not. Images were taken at MNI [−27, −27, −30]. (B) Visual representation of the relationship between LLP – L parahippocampus connectivity and tobacco use frequency. (C) On the contralateral side, LLP – R parahippocampus connectivity reached trend-level association with tobacco use frequency.

Although these clusters were lateralized, the thresholded connectivity maps showed clusters in bilateral hippocampus and parahippocampus (Figures 1A, 2A). To investigate the laterality of the connectivity-tobacco association, we tested the association between tobacco use and LLP connectivity to regions contralateral to brainwide discovery clusters (L hippocampus, MNI [−30,-12, −27]; R parahippocampus, MNI [+27, −27, −27]). LLP – L hippocampus connectivity was significantly associated with tobacco use frequency (estimate=0.035, SE=0.016, t=2.175, p=0.031, Figure 1C). LLP – R parahippocampus connectivity reached trend-level association with tobacco use frequency (Figure 2B).

### CHR-P Diagnosis Enhanced the Association between LLP – R Hippocampus Connectivity and Tobacco Use Frequency

LLP - R hippocampus connectivity was higher in current tobacco users compared to nonusers (t=-3.5466, df=101.88, p=0.0006; Figure 3A), and higher in CHR than control (t=-2.8651, df=279.47, p=0.0049; Figure 3B). We did not observe significant tobacco x diagnosis interactive effect on LLP - R hippocampus connectivity across the entire sample. However, among those with current tobacco use, there was a significant tobacco x diagnosis interaction on LLP - R hippocampus connectivity (estimate=0.306, SE=0.149, t=2.051, p=0.045; Supplemental Figure 1), when modeling tobacco use as heavy use (>10 cigarettes/day) or light use (<10 cigarettes/day), and controlling for cannabis use. Post-hoc analysis using estimated marginal means did not reveal significant differences of predicted connectivity between groups, though there was trend-level difference between connectivity of CHR-heavy use and control-heavy use (p=0.071).

**Figure 3.**
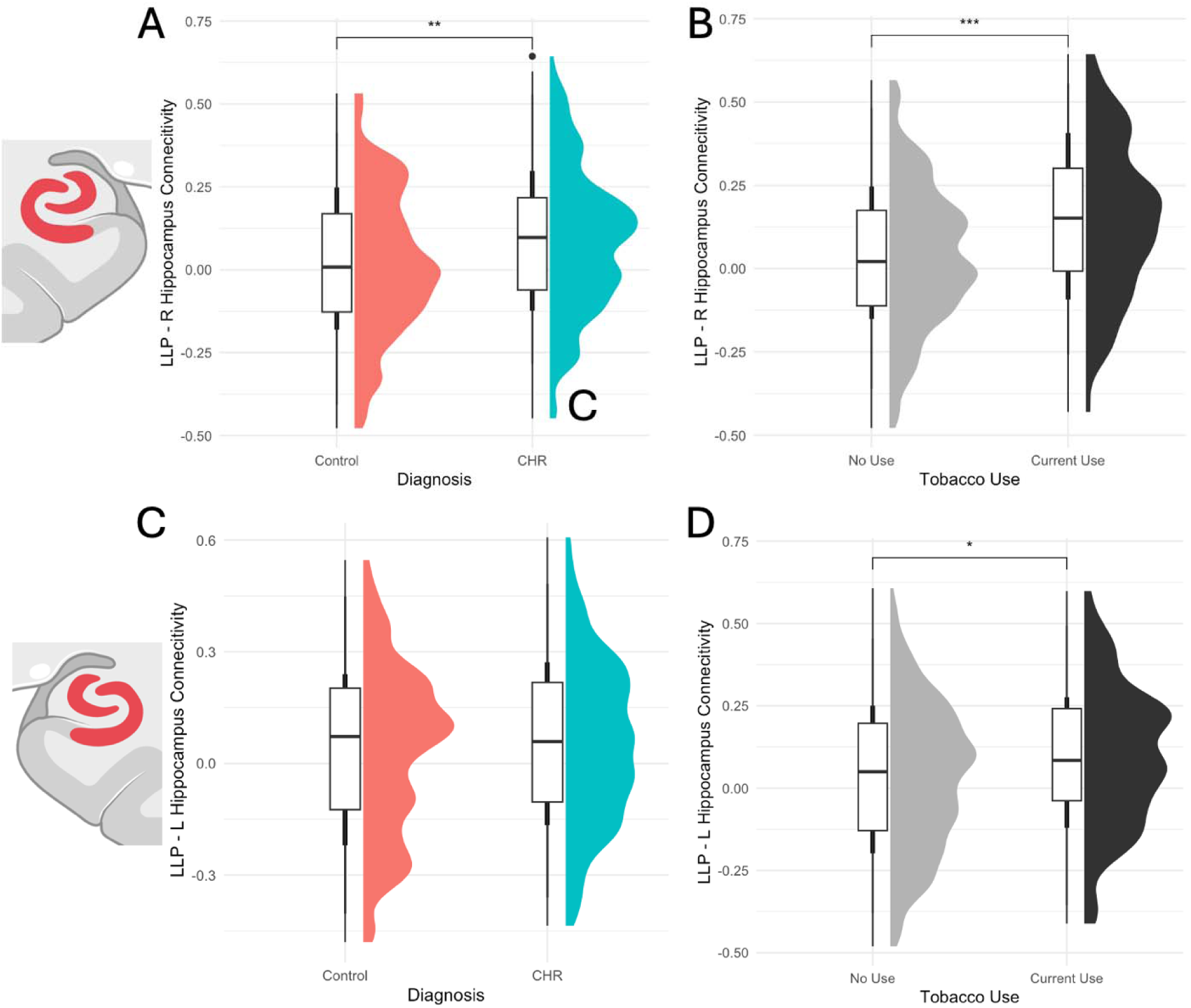
Comparison of LLP-Hippocampus Connectivity by Diagnosis and by Tobacco Use. LLP - R hippocampus connectivity was (A) higher in current tobacco users compared to nonusers (t=-3.5466, df=101.88, p=0.0006), and (B) higher in CHR than control (t=-2.8651, df=279.47, p=0.0049).

### LLP Connectivity to L Hippocampus and Bilateral Parahippocampus Differed by Tobacco Use but not by CHR-P Diagnosis

Compared to nonusers, current tobacco users had higher connectivity from LLP to L hippocampus (t=-2.108, df=111.59, p=0.037; Figure 3D), R parahippocampus (t=-2.015, df=105.88, p=0.037; Figure 4B), and L parahippocampus (t=-2.725, df=96.561, p=0.0076; Figure 4D). None of the three connectivity edges differed by CHR-P diagnosis (p>0.1; Figures 3C, 4A, 4C). We did not observe a tobacco x diagnosis interaction on connectivity for these edges (p>0.1).

**Figure 4.**
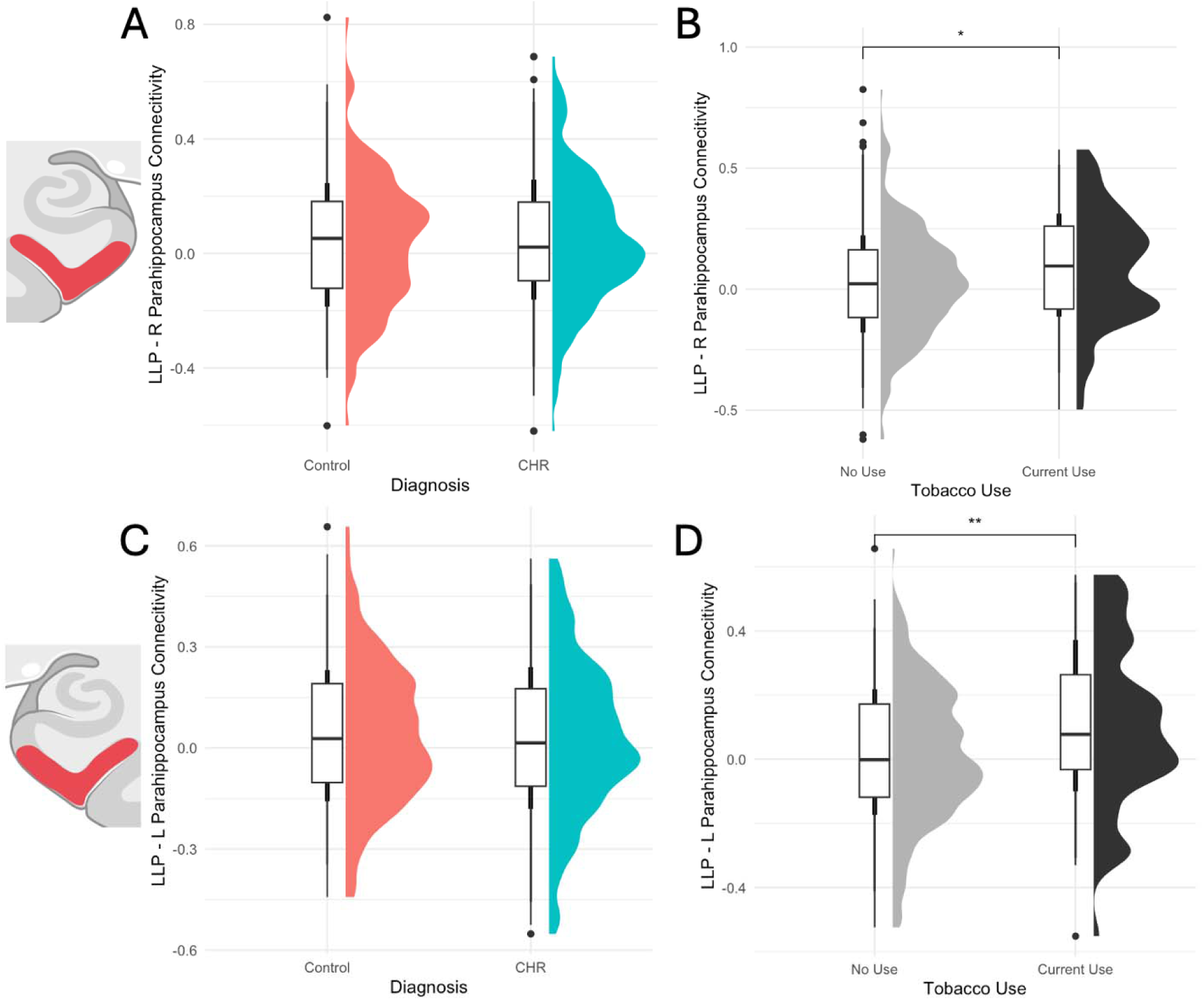
Comparison of LLP-Parahippocampus Connectivity by Diagnosis and by Tobacco Use. Bilateral parahippocampus connectivity to LLP did not differ by CHR-P diagnosis (A,C). Compared to nonusers, current tobacco users had (B) higher LLP - R parahippocampu connectivity (t=-2.015, df=105.88, p=0.037) and (D) higher LLP - L parahippocampus connectivity (t=-2.725, df=96.561, p=0.0076).

## Discussion

Using the NAPLS2 neuroimaging and substance use data, we investigated the relationship between tobacco use and resting state functional connectivity of a parietal DMN node in CHR-P. We performed an unrestricted, brainwide analysis by regressing connectivity against tobacco use frequency. We found that more frequent tobacco use is associated with stronger parietal DMN connectivity to bilateral hippocampus. We also observed a diagnosis-by-connectivity interaction among current tobacco users, where higher tobacco use predicted hyperconnectivity between parietal DMN and R hippocampus only in CHR, and hypoconnectivity in HC.

We observed higher connectivity between parietal DMN and R hippocampus in CHR-P compared to control individuals. Hippocampal involvement in this population has been well-characterized. Hippocampal hyperactivity has been consistently observed in CHR-P^22–24^, and the degree of hyperperfusion correlated with psychotic symptom severity^22^. Beyond exhibiting hippocampal hyperactivity, CHR-P individuals also demonstrated altered hippocampal connectivity to cortical regions, including reduced connectivity between hippocampus and frontal regions^25,26^, and association between hippocampus-brainwide dysconnectivity and symptom severity^27^.

While there is limited characterization of hippocampal-DMN connectivity in CHR-P, this connectivity has been studied in SZ. Kragulijac et al observed decreased hippocampal-precuneus connectivity in unmedicated patients with chronic SZ^28^. Among medicated SZ patients, Nelson et al reported increased hippocampal connectivity with precuneus and lingual gyrus in SZ^29^, while McHugo et al did not report differences in hippocampal-DMN connectivity^30^. However,

McHugo et al proposed the possibility that instead of exhibiting hippocampal dysconnectivity to the entire DMN, only certain DMN subregions are altered in psychotic illness. Our results indicated that hippocampus – parietal DMN dysconnectivity emerges at an early stage of the illness. Future research can characterize how this connectivity edge evolves through illness progression to investigate its validity as a possible biomarker or an intervention target.

We hypothesized that higher tobacco use would correlate with higher intra-DMN connectivity. While we did not find hyperconnectivity within canonical regions of the DMN, some have conceptualized the hippocampus as a part of the DMN^31^. The hippocampus is both structurally and functionally connected to the DMN. Structurally, the posterior hippocampus is connected via the fornix pathway to thalamus and retrosplenial cortex, both are deep DMN regions^32^. Human resting-state fMRI studies also reported functional connectivity between posterior hippocampus and posterior DMN regions, including the posterior cingulate and the precuneus^32–36^. Both resting-state and task-deactivation analyses have reported medial temporal lobe involvement in the DMN^36–38^. Our finding is consistent with this line of evidence supporting the connectivity between the hippocampus and the DMN. Further, we extended the current knowledge of hippocampal connectivity to deep DMN regions, showing that the hippocampus is also functionally connected to superficial DMN regions, particularly in the presence of current tobacco use.

We found a diagnosis-by-tobacco interaction on connectivity, where frequency of tobacco use differentially impacted hippocampal – parietal DMN connectivity in CHR-P compared to control individuals. This diagnosis-specific finding is consistent with current knowledge on hippocampal response to nicotine signaling in CHR-P. Nicotine directly activates nicotinic-acetylcholine receptors (nAChR) in the hippocampus^39^. Different subregions of the hippocampus have differential nAChR expression, but they are found on nearly every synaptic level^40^. Nicotine binding to nAChR causes neuron depolarization to activate cell-dependent downstream effects, and indirectly alter nAChR expression^40^. The latter can disrupt cholinergic signaling, which leaves widespread and longstanding impacts on cell functioning, hippocampus morphology, and regulation of other neurotransmitters. These impacts are especially significant during periods of rapid hippocampus remodeling, such as in adolescence^40^ and in early/prodromal stage of psychotic illnesses^41^. CHR-P individuals, who are often adolescents, are thus even more vulnerable to developing persistent nicotine-induced disruptions compared to general adolescents. It is possible that early tobacco exposure compounded the downstream effects of cholinergic signaling disruption from the psychosis illness in CHR-P. Future research can study the longitudinal relationship between tobacco/nicotine exposure, neurotransmitter signaling, and brain connectivity across stages of illness development.

Considering CHR-P’s vulnerability to developing persistent cellular and biochemical changes from nicotine exposure, nicotine cessation interventions are particularly important in this population. Recent clinical trials have assessed the effect of nAChR agonists on psychotic symptoms and cognitive performance. One such compound demonstrated some positive impact on cognition^42^ and negative symptom severity^43^ in patients with schizophrenia compared to placebo. However, the intrinsic quick desensitization properties of the receptor may limit the duration of efficacy of pharmacologic agents, limiting their utility as a treatment^44^. Another possible intervention is through transcranial magnetic stimulation (TMS), a form of noninvasive neuromodulation. The putative mechanism of TMS is by directly impacting the node of application, thereby indirectly impacting activity of neural networks functionally connected to this region^45^. Although TMS cannot directly modulate the hippocampus due to its deep location in the brain, it is possible to affect the hippocampus by modulating its functionally connected regions, such as the DMN^46^. Previous works showed that TMS applied to the left lateral parietal node of DMN (LLP) successfully modulated both DMN connectivity and nicotine craving in SZ^47^, but not in smokers without psychosis^48^. A recent clinical trial reported that TMS improved cognitive performance and changed hippocampus connectivity to posterior DMN in individuals with mild Alzheimer’s Disease^49^. Our hippocampal – parietal DMN connectivity association with tobacco use in CHR-P builds the theoretical basis in potentially modulating hippocampal connectivity through DMN regions.

This study has several strengths, including use of a brainwide analysis to discover novel connectivity edges associated with tobacco use, instead of limiting our scope to established resting-state functioning connectivity networks. Our analysis was well-powered by a large sample recruited from multiple study sites across North America. Substance use assessment was relatively comprehensive for this population, including quantifying tobacco use frequency instead of binary assessment of use or no-use, assessment of other substance use, and having a specific time window (past 30 days) of substance exposure. Several limitations exist as well. The major limitation is the difficulty in disentangling cause from effect using a cross-sectional study design. Future studies should include longitudinal neuroimaging to capture longitudinal brain changes under the joint influence of tobacco use, brain development, and progression or recovery in the CHR state. Additionally, this sample has significantly fewer number of people who use tobacco in the control group compared to the CHR-P group, though we accounted for this heteroscedastic sample distribution using robust standard error. Moreover, regular 3T resting-state fMRI is not the best imaging protocol for capturing details of the medial temporal lobe. We did not use a hippocampus-specific imaging protocol because we did not have an a priori hypothesis on hippocampal involvement.

In conclusion, we found that more frequent tobacco use is associated with stronger parietal DMN connectivity to bilateral hippocampus in NAPLS2, and CHR-P diagnosis influenced the relationship between tobacco use and connectivity. At an early stage of psychosis illness development, altered hippocampus – parietal DMN connectivity is present and linked to tobacco use. Our results add evidence to support the role of DMN connectivity in the high prevalence of tobacco use in both CHR-P and psychotic-spectrum disorders.

## Data Availability

All data produced in the present study are available upon reasonable request to the authors.

## Acknowledgments

This work was supported by National Institutes of Health (NIH) grants U01 MH066134 to Dr. Addington, P50 MH066286 to Dr. Bearden, U01 MH081944 to Dr. Cadenhead, U01 MH081902 to Dr. Cannon, U01 MH081857 to Dr. Cornblatt, R01 MH076989 to Dr. Mathalon, U01 MH066069 to Dr. Perkins, U01 MH081928 to Dr. Stone, U01 MH081988 to Dr. Walker, U01 MH82022 to Dr. Woods, R01 MH116170 to Dr. Brady, K23DA059690 to Dr. Ward, and Vanderbilt Medical Scholars Fellowship Program to Ms. Bai.

## Competing Interests

The authors have no competing interests to disclose.

Note: Barbara Cornblatt and Larry Seidman passed away tragically before submission of this manuscript. Their colleagues wish to honor their contributions to the work posthumously.

