## Supplement for "Tobacco Use is Related to Parietal-Hippocampal Connectivity in People at Clinical High Risk for Psychosis"

**Methods**

***Participants***

The North American Prodrome Longitudinal Study (NAPLS2) is a longitudinal case-control study studying the psychosis prodrome across 8 sites in North America. Baseline neurocognitive functioning data and resting-state fMRI were collected from January 2009 to April 2013. A total of 211 CHR and 132 control participants completed baseline were enrolled and had complete neuroimaging and cognitive data. The complete methods and clinical assessments of the NAPLS2 study are described in detail elsewhere^1^. The study protocols were approved the institutional review boards of all sites, including Beth Israel Deaconess Medical Center, Boston, Massachusetts; Emory University, Atlanta, Georgia; University of Calgary, Alberta, Canada; University of California, Los Angeles; University of California, San Diego; The University of North Carolina at Chapel Hill; Yale University, New Haven, Connecticut; and Zucker Hillside Hospital, Queens, New York. All participants provided written informed consent.

***Inclusion and Exclusion Criteria***

CHR individuals were included if they met the Criteria of Prodromal Syndromes (COPS), based on the Structured Interview for Prodromal Symptoms (SIPS). If individuals were younger than 19 years, they were included based on criteria for schizotypal personality disorder or COPS. Anyone with a lifetime Axis I psychotic disorder, estimated IQ less than 70 on both measures of IQ, a central nervous system disorder, or DSM-IV substance dependence in the past 6 months was excluded. Other nonpsychotic DSM-IV disorders were not exclusionary (e.g. depression, substance use disorders) unless they clearly caused or better explained prodromal symptoms. CHR individuals were permitted to take antipsychotic medication as long as they had not developed any psychotic symptoms prior to initiating medication. HCs were not permitted to meet any prodromal criteria, have a history of a psychotic or cluster A personality disorder, or have a family history of a psychotic disorder in a first-degree relative.

***Clinical Measures***

The Structured Clinical Interview for DSM was used to exclude psychosis and to identify DSM-IV Axis I or cluster A personality disorders. Conversion to psychosis was determined if a subject met the SIPS Presence of Psychotic Symptoms criteria.

***MRI Data Acquisition***

All participants underwent a 5-min eyes-open resting-state scan (154 whole-brain volumes), where subjects were asked to lay still in the scanner, relax, gaze at a fixation cross, and not engage in any particular mental activity^2^. After the scan, investigators confirmed with the participants that they had not fallen asleep in the scanner. Data were acquired from eight 3T MR scanners with identical fMRI protocols. Siemens scanners were used at Emory, Harvard, UCLA, UNC and Yale, and GE scanners were used at Calgary, UCSD and ZHH. Functional images were collected using gradient-recalled-echo echo-planar imaging (GRE-EPI) sequences: TR/TE 2000/30 ms, 77 degree flip angle, 30 4-mm slices, 1-mm gap, 220-mm FOV. In addition, we also acquired high-resolution T1-weighted images for each participant with the following sequence: 1) Siemens scanners: magnetization prepared rapid acquisition gradient-echo (MPRAGE) sequence with 256 mm x 240 mm x 176 mm FOV, TR/TE 2300/2.91 ms, 9 degree flip angle; 2) GE scanners: 3 spoiled gradient recalled-echo (SPGR) sequence with 260 mm FOV, TR/TE 7.0/minimum full ms, 8 degree flip angle.

NAPLS2 imaging procedures and assessments were designed in a highly coordinated manner. All procedures were harmonized from conception and design of the study prior to data collection^1^. A standardized set of scan parameters was implemented across all sites/scanners. Quality assurance methods included the use of a common structural phantom scanned at each site.

***MRI Data Processing***

All analyses were preprocessed using the DPABI toolbox (Data Processing and Analysis for Brain Imaging; http:// rfmri.org/dpabi)^3^. As a quality control metric, data from any participant whose scans exceeded motion thresholds (3 mm translation or 3° rotation) were discarded. Individual time points with framewise displacement 0.2 mm were removed via scrubbing^4^, and scans with 50% of volumes removed for framewise displacement were discarded. All data were preprocessed to remove motion (24-parameter), CSF signals, white matter signals, global signal, and overall linear trend. A bandpass filter was applied (0.01–0.08 Hz). Data were normalized using the DARTEL toolbox into Montreal Neurological Institute (MNI) space and smoothed with an 8-mm full-width half-maximum kernel. Analyses were conducted in a gray matter mask defined within the group. Data were resampled into 4mm isotropic resolution prior to further analyses.

**Supplemental Figure 1**

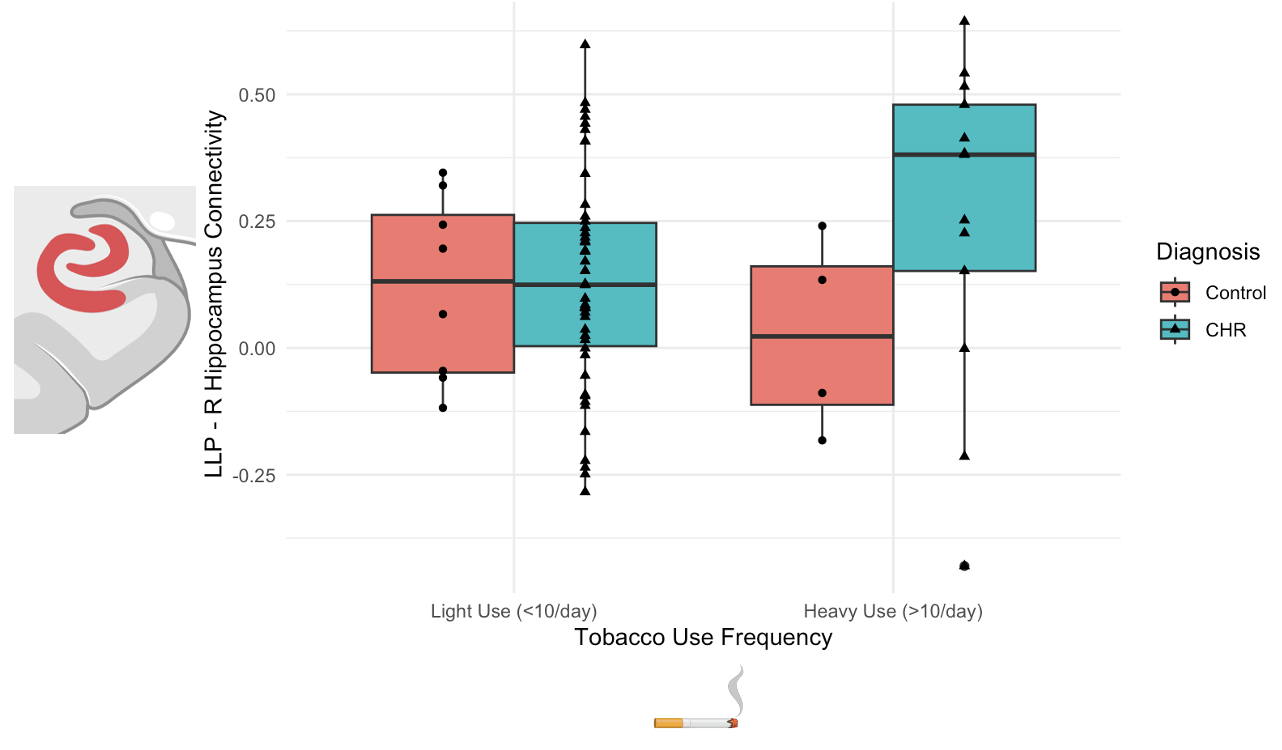
